# Genetic susceptibility of Saudi Population to Hepatitis B Virus (HBV) infection and the predicted functional consequences

**DOI:** 10.1101/2025.03.16.25323998

**Authors:** Saira Sarfraz Khalid, Khalid Alswat

**Affiliations:** Liver Disease Research Centre, King Saud University, Riyadh, Saudi Arabia; Liver Disease Research Centre, College of Medicine, King Saud University, Riyadh, Saudi Arabia

**Author notes:** Corresponding Author: Saira S Khalid, Liver Disease Research Centre, King Saud University, PO Box 2925 (59), Riyadh 11461, Saudi Arabia.

**Keywords:** Hepatitis B virus, single nucleotide polymorphism, Saudi Arabia, genetic variations

## Abstract

Chronic Hepatitis B virus (HBV) infection poses a global public health challenge, for which an effective cure remains elusive. A substantial amount of data has shown that single nucleotide polymorphisms (SNPs) within host genes can affect the regulation and expression of proteins, thereby influencing the susceptibility to HBV infection as well as disease progression and response to treatment. HBV-related SNPs have been identified in the population of Saudi Arabia, however, there is a lack of in-depth characterization of the translational and functional impact of these SNPs. This article aims to analyze the SNPs significantly associated with HBV-associated complications in the Saudi population, predict their functional impact using bioinformatic tools and propose future projections for HBV research in Saudi Arabia. The findings of these genetic studies are likely to pave the way for developing more effective preventive and therapeutic interventions by personalizing the management of HBV infection.

## Introduction

Despite the advancement in treatment and the implementation of a vaccine program, as well as the global hepatitis elimination targets set by the World Health Organization, an estimated 254 million people worldwide are still affected by the hepatitis B virus (HBV) (1). These individuals require monitoring of viral activity, and disease progression along with appropriate treatment when indicated to minimize their risk of developing liver complications like cirrhosis and hepatocellular carcinoma (HCC) (2) thereby posing significant challenges to public health. In addition to that, a substantial number of infected individuals remain undiagnosed due to the silent nature of the disease, leading to potential ongoing transmission of the infection (1). In Saudi Arabia, the prevalence of HBV infection is estimated to be around 1.3 %, marking it as a region of low endemicity. However, the control and management of HBV remain a signified challenge for healthcare systems with an increase in HBV-related HCC cases and other comorbidities (3, 4). Regardless of the implementation of mandatory pregnancy and pre-marital screening policies in Saudi Arabia, there is a lack of awareness about disease transmission. Furthermore, social stigmatization related to HBV disease increases the disease burden in the country (5). Non-adherence to HBV treatment, especially in asymptomatic patients, also hinders its management and care (5). These factors have urged the hunt for reliable biomarkers that could predict disease and treatment outcomes.

The literature review shows that host genetic factors can impact the clinical outcome and treatment response in patients with chronic hepatitis B (6, 7), suggesting their potential use as predictive or prognostic biomarkers. The most commonly studied genetic factors are single nucleotide polymorphisms (SNPs) or genetic variations, which can affect gene expression and translation processes, thereby impacting disease development (8). SNPs can be found within the coding region of a gene, leading to either a synonymous (silent) or nonsynonymous change. Nonsynonymous SNPs (nsSNPs) result in the substitution of amino acids in the respective protein, potentially impacting its structure, function, stability, and solubility (9). On the other hand, synonymous SNPs do not change the nature of amino acids but may affect translation rates or mRNA half-life. In addition, many SNPs were found in the non-coding regions of the genome, which impact transcriptional activity or gene expression and processing (9). Sometimes, a set of SNPs along a single chromosome tends to be inherited together, forming haplotypes that can collectively affect the same gene (10).

The identification of disease-related SNPs can be achieved through DNA sequencing techniques or genome-wide association studies (GWAS). Numerous studies have been conducted on samples from patients with Hepatitis B to identify specific SNPs that are associated with susceptibility to HBV, as well as disease progression and response to treatment (Reviewed in (11, 12)). The distribution and importance of these SNPs differ among various populations and ethnic groups, prompting genetic investigations in diverse ethnic populations to address these disparities. Although genome studies in the HBV-infected population of Saudi Arabia have uncovered both known and novel SNPs associated with HBV infection and disease advancement, these findings have yet to be further examined or implicated in subsequent research. This article aims to investigate the coding and non-coding HBV-associated SNPs identified in the Saudi Arabian population and predict their translational or functional role using bioinformatics tools. It will also examine how these SNPs impact host defense pathways and their role in contributing to HBV susceptibility within this population. Lastly, we propose how this knowledge can guide future research and be incorporated into a multicentre platform.

### Search method and analysis strategy

This study is a review based solely on previously published research. No new data involving human participants or animals were collected, and no identifiable patient information was used. Therefore, ethical approval was not required for this study.

The genetic studies related to HBV-associated disease in the Saudi Arabian population were searched on Pubmed and Scopus using the keywords “Hepatitis B” or “hepatitis B virus”, “genetic studies”, “genome-wide association study”, “single nucleotide polymorphism” and “Saudi Arabia”. A total of nine publications were found with significant findings, that were conducted by research teams with more or less similar members or authors, indicating their shared interest in the topic. These nine publications studied a total of thirty-eight SNPs belonging to eleven genes associated with the antigen presentation pathway and immune pathways (receptors and cytokines). The primary information collected from each study includes sample size, SNPs studied, major and minor alleles and their frequencies, SNPs with significant association with HBV, and their p-value and odds ratio (O.R.) (Supplementary Table 1).

Out of the thirty eight SNPs analyzed in the nine selected studies, twenty-three were significantly associated with HBV infection and were uploaded in SNPNexus (https://www.snp-nexus.org/v4/). SNPnexus is a web-based annotation tool that provides genomic coordinates for the queried SNPs regarding their chromosomal position and a comprehensive overview of potential functional consequences of SNPs in coding regions such as synonymous or non-synonymous and deleterious or tolerated effects on protein function (13). Moreover, for SNPs in the non-coding regions, it provides regulatory impact, i.e. whether the SNP disrupts transcription factor-binding sites (TFBS) or potentially alters the transcriptional and post-transcriptional regulation of gene expression. It also provides scores for the non-coding SNPs using CADD (Combined Annotation-Dependent Depletion) algorithm, which predicts the potential deleterious impact of genetic variants in the form of values (14). A value of above 20 is considered deleterious while less then 20 benign. To understand the effect of the non-synonymous SNPs (nsSNPs) on the protein structure, we used ProtVar (https://www.ebi.ac.uk/ProtVar) and HOPE (https://www3.cmbi.umcn.nl/hope) web servers that identify the structural effect of the point mutations in protein sequences. The UniProt-Accession Codes for NOD2 (Q9HC29), MD2 (Q9Y6Y9), CXCR1 (P25024), and IL37 (Q9NZH6) were used individually as inputs for the analysis. To predict the relation of nsSNPs of NOD2 with pathogenecity we use Mutpred2 serevr (http://mutpred.mutdb.org/index.html) that can predict if detailed molecular target and affected mechanism with a score ranging from 0 to 1. The closer the Metpred score to 1, the higher the chance of the substitution to alter protein function. The schematic diagram for the analysis is shown in Figure 1.

**Figure 1.**
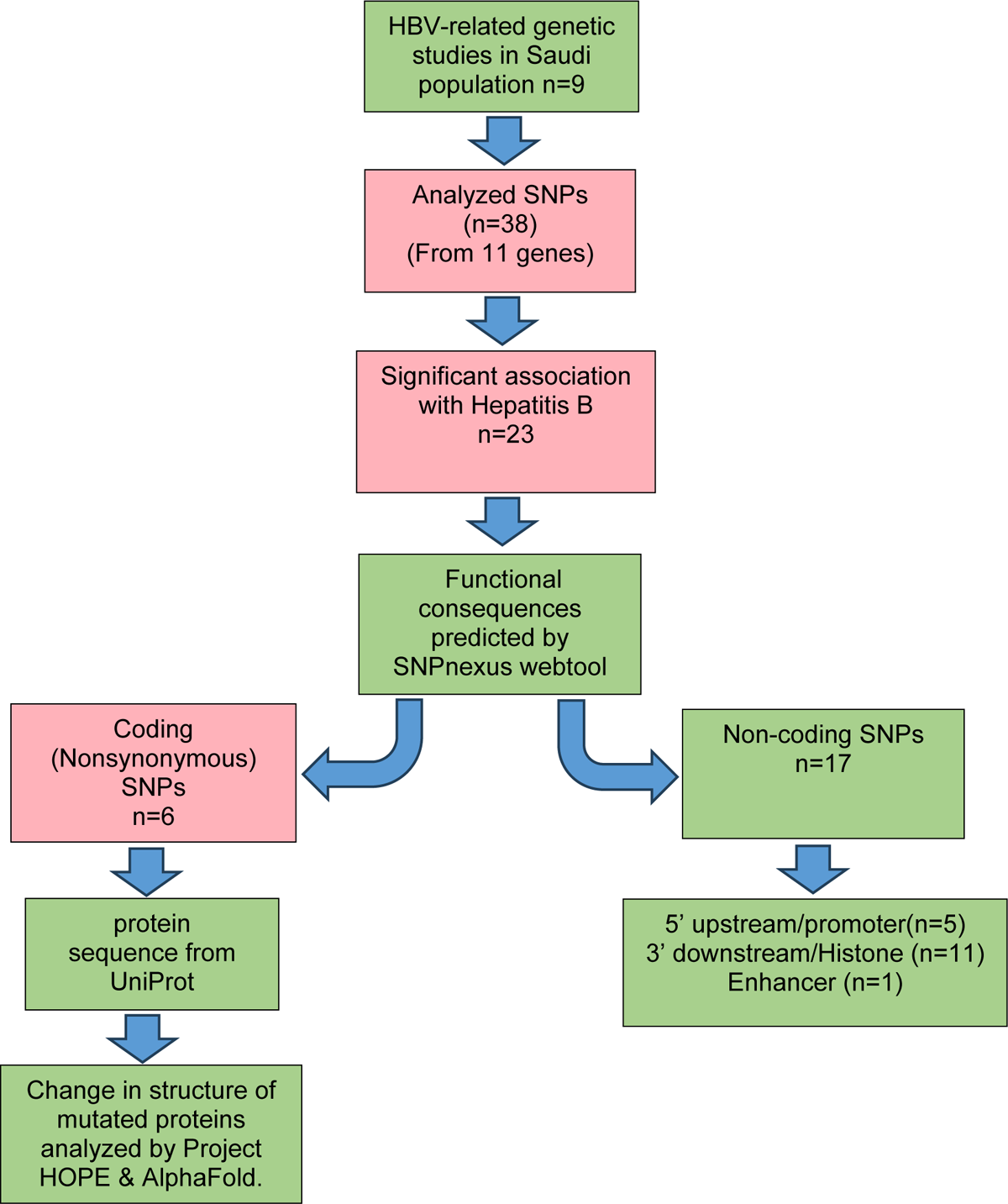
Schematic diagram summarizing the study. The analysis of HBV-associated host SNPs using bioinformatic tools to predict their functional consequences.

### Functional annotation of SNPs identified from Genetic studies in the Saudi Arabian population

The genetic studies conducted in the Saudi Arabian population were carried out through a collaboration of multiple centers. The distribution of SNPs also known as genetic variations was compared among different groups, including healthy controls, clearance group, inactive HBV infection, active HBV infection, HBV-associated cirrhosis, and HCC. The clearance group was defined as the subjects with spontaneous HBV clearance having hepatitis B surface antigen (HBsAg)-negative, Anti-HBV core antibody positive, and HBV DNA negative. The inactive HBV group was defined as subjects with HBsAg-positive and hepatitis B e antigen (HBeAg)-negative with persistently normal alanine aminotransferase (ALT) and HBV DNA persistently <2000 IU/mL, whereas active HBV group was defined as subjects with either HBeAg-negative or HBeAg-positive, with elevated ALT levels and HBV DNA ≥2000 IU/ml (15). The sample size of the hepatitis B patients in all studies was over 600, while that of the healthy control and clearance groups was over 500. They performed a single GWAS, while the rest of the studies used DNA sequencing techniques. The SNPs selected in the DNA sequencing techniques belong to genes related to immune response pathways and inflammatory mediators. According to the authors, the selection of these SNPs was meticulously based on published literature or the crucial importance of the roles of the genes in the outcome of HBV infection. Of the twenty-three significantly associated SNPs, six SNPs (rs2066845, rs2066844, rs6472812, rs11466004, rs2234671, and rs28947200) were located in coding regions of four genes (NOD2, MD2, CXCR1, and IL37). The remaining 17 SNPs were in non-coding regions, which included 5’ untranslated regions (UTR) or promoter regions and 3’ untranslated regions and had predicted functions as histone regulatory elements (Table 1).

**Table 1.** Genomic coordinates and predicted consequences of HBV-related SNPs by SNPnexus. FDX1=Ferredoxin 1, *HLA-DQB=Human Leukocyte Antigen DQ region Beta chain, HLA-DPAor B=Human Leukocyte Antigen DP region alpha or beta chain, MICA=MHC class I Chain A, TLR3 =Toll like receptor 3,* NOD2=Nucleotide binding oligomerization domain 2, MD2= myeloid differentiation protein 2, CXCR1 = Chemkine C-X-C motif receptor 1, IFNL3 or 4 = Interferon Lambda 3 or 4, IL37 = Interleukin 37.

| SNP-allele | chromosome | Nearest Gene | Annotation | Predicted function | CADD score |
| --- | --- | --- | --- | --- | --- |
| rs2724432 | chr11 | FDX 1 | Regulatory element | Histone | 8.2 |
| rs2856718 | chr6 | HLA-DQB | Intronic | Promoter flanking region | 8.611 |
| rs9275572 | chr6 | HLA-DQ | Intronic | Histone/transcription factor | 5.595 |
| rs3077 | chr6 | HLA-DPA | Non-coding, 3utr | Histone/Transcription factor | 9.602 |
| rs9277535 | chr6 | HLA-DPB | 3downstream,/3UTR |  | 6.14 |
| rs2596542 | chr6 | MICA | 5 upstream | Promoter | 2.054 |
| rs1879026 | chr4 | TLR-3 | Intronic | Promoter flanking region | 0.02 |
| **rs2066845** | chr16 | NOD-2 | Coding, non-synonymous | Deleterious | 27.4 |
| **rs2066844** | chr16 |  | Coding, nonsynonymous | Deleterious | 23.6 |
| **rs6472812** | chr8 | MD-2 | coding non-synonymous | Tolerated | 11.2 |
| **rs11466004** |  |  | Coding, nonsynonymous | Tolerated | 13.4 |
| **rs2234671** | chr2 | CXCR1 | Coding non-synonymous | Tolerated | 2.9 |
| rs8105790 | chr19 | IFNL3 | 3downstream | Histone | 2.1 |
| rs12979860 | chr19 | INFL4 | Intronic, | Histone/Transcription factor. | 1.7 |
| rs12980275 | chr19 | IFNL3 | Intronic | Histone | 0.689 |
| rs2723175 | chr2 | IL37 | Intronic, upstream | Histone | 0.63 |
| rs2708973 | chr2 | IL37 | Upstream | Enhancer. | 4.56 |
| rs2723176 |  |  | Intronic, 5upstream | Promoter flanking region. | 3.2 |
| rs2723186 | chr2 | IL37 | Intronic | Histone | 0.15 |
| **rs28947200** | chr2 | IL37 | Coding non-synonymous | Deleterious | 19.1 |
| rs4364030 | chr2 | IL37 | Upstream | Histone | 1.0 |
| rs4392270 | chr2 | IL37 | downstream | CTCF binding site | 1.78 |
| rs4849133 | chr2 | IL37 | downstream | Promoter flanking region/Transcription factor | 2.3 |

In the following paragraphs, the twenty-three significantly associated SNPs were categorized according to their gene’s function. The involvement of relevant genes in hepatitis B disease was discussed, along with the findings of bioinformatic tools regarding their functional impact. Additionally, the SNPs were categorized based on their association either with HBV persistence or clearance (Table 2).

**Table 2.** Genetic risk alleles associated with Hepatitis B clearance, suceptibility, active disease and cirrhosis or HCC FDX1=Ferredoxin 1, *HLA-DQ=Human Leukocyte Antigen DQ region, HLA-DP=Human Leukocyte Antigen DP region, MICA=MHC class I Chain A, TLR3 =Toll like receptor 3,* NOD2=Nucleotide binding oligomerization domain 2, MD2= myeloid differentiation protein 2, CXCR1 = Chemkine C-X-C motif receptor 1, IL28 = Interlekukin 28, IL37 = Interleukin 37.

| **HBV Clearance / Protection against HBV** | **HBV susceptibility** | **Active HBV (compared to inactive)** | **HBV-cirr/HCC (compared to active)** |
| --- | --- | --- | --- |
|  |  |  | **FDX 1**<br>rs2724432 |
| ***HLA-DQ***<br>rs9275572-A | rs2856718-A<br>***HLA-DP***<br>rs3077-G<br>rs9277535-G |  |  |
|  |  | ***MICA***<br>rs2596542-T |  |
| ***TLR3***<br>rs1879026-T |  |  |  |
| ***NOD2*** | rs2066844-T<br>rs2066845-GC |  | rs2066845-CC |
| ***MD2***<br>rs6472812-A<br>rs11466004-CT |  |  |  |
| ***CXCR-1*** |  | rs2234671-C | rs2234671-C |
| ***IL28***<br>rs8105790-C<br>rs12980275-G | rs12979860-T | rs12979860-T | rs12979860-T |
| ***IL37***<br>rs2723176-A<br>rs2723186-AG<br>rs4364030-G<br>rs4392270-AG | rs2723175-A<br>rs2708973-A<br>rs28947200-Tor<br>CT | rs4392270-A<br>rs4849133-CT |  |

### GWAS identified SNP or risk allele, Ferredoxin 1 (FDX1)

The first GWAS performed in HBV-infected Saudi subjects identified the SNP rs2724432-T allele to be strongly associated with cirrhosis and HCC when compared with inactive-HBV group (16). This SNP is located upstream of the Ferredoxin 1 (FDX1) gene, a small electron transfer protein involved in bile and cholesterol metabolisms (17). There is no subsequent study by any group to analyze the biological impact of this SNP or FDX1 gene in hepatitis B associated disease. However, a recent research identified an imperative role for FDX1 in regulating HCC progression, by showing that reduced expression of FDX1 increases oxidative stress and mitochondrial degradation which facilitates HCC proliferation, invasion and metastasis (18). These observation support the association found between FDX1 and HBV-mediated cirrhosis or HCC in Qahtani et al study. The SNPNexus analysis classified this SNP as a histone-related regulatory element with a CADD score of 8.2. Although this score is lower than the typical threshold for deleterious variants (such as a CADD score of twenty or more), it is still meaningful, especially in its potential impact on gene transcription, which can result in altering gene expression. Histones are proteins that package and organize DNA into nucleosomes, enabling DNA chromatin to relax or compress, thereby regulating gene expression (19). SNPs in these regions may influence chromatin structure or gene expression by affecting histone modification like acetylation or methylation. Consequently, the rs2724432 risk allele could affect the expression levels of FDX1, and further studies are required to determine whether its association with HBV-mediated cirrhosis or HCC is substantial. Since FDX 1 is not a secreted protein, its levels can be examined through immunostaining with FDX1 antibodies on archival liver biopsies from patients at various stages of chronic hepatitis B. This approach could offer insights into the role of this protein in disease progression.

### Immune system-related genes

The clinical outcome of HBV infection depends on the balance between the virus’s ability to persist and the host’s ability to clear it. This determines whether the infection will be acute, chronic, or progress to more severe liver disease. During HBV infection, various immune cells such as dendritic cell, macrophages and cytotoxic T cell are involved in recognizing the viral particels through antigen presenting molecules and pathogen recognition receptors (PRR). These receptors activate other immune cells to eliminate infected cell or release cytokines and interferons to prevent HBV replication. Therefore, most SNPs selected in HBV-related host genetic studies were related to different parts of the immune system.

### Antigen Presenting molecule (HLA-DQ, HLA-DP and MICA)

The human leukocyte antigen (HLA) molecules are equivalent to Major Histocompatibility complexes (MHC) that bind foreign antigens, present them to T cells (CD4/CD8), and initiate an immune response. There are two classes of HLA molecules: class I, which binds endogenous antigens and presents them to CD8 (T) cells, and class II which binds to exogenous antigens and presents them to CD4 T cells (20). The GWAS and DNA sequencing techniques, performed in a variety of populations, have identified various SNPs in Class II HLA molecules, spedifically in DQ and DP regions that are linked to susceptibility to HBV (Reviewed in (6, 12). In the Saudi cohort conducted by Al-Qahtani and colleagues, they found that HLA-DQ rs9275572-A alleles were significantly associated with protection against HBV, whereas HLA-DQ rs2856718-A, along with HLA-DP rs3077-G and rs9277535-G polymorphism, were significantly associated with a higher risk of HBV infection. Their haplotype analysis showed that the presence of the haplotypes GG of rs3077 and 9277535 was significantly associated with HBV susceptibility (15).

SNPnexus analysis shows predicted function of rs2856718 to be related to the promoter region of HLA-DQ beta chain with a CADD score of 8.6. This suggest that the presence of risk allele could influence mRNA expression of HLA-DQ through modifications in promoter activity and transcription factor binding (21). In contrast, rs3077 and rs9277535 variants are located in the 3’UTR, and have CADD scores of 9.6 and 6.1, respectively. The SNPs in 3’UTR can have functional impacts on mRNA stability, localization, and translation efficiency (22). A previous study has shown that risk alleles of rs3077 and 9277535 were associated with decreased mRNA levels of HLA-DPA1 and HLA-DPB1 in normal liver samples (23). Based on these findings, it can be concluded that HLA-related SNPs, such as rs3077 and rs9277535, may reduce the expression of their respective proteins, thereby impairing the immune system’s ability to recognize and respond to HBV infection. This impaired immune recognition could increase susceptibility to chronic HBV infection (Figure 2a). Future studies should focus on validating these findings by quantitatively assessing the mRNA and protein expression levels of HLA-DQ and DP in Saudi Hepatitis B patients with risk alleles. Additionally, functional assays should be conducted to evaluate the downstream immune response particularly the activation of APC and T cells.

**Figure 2.**
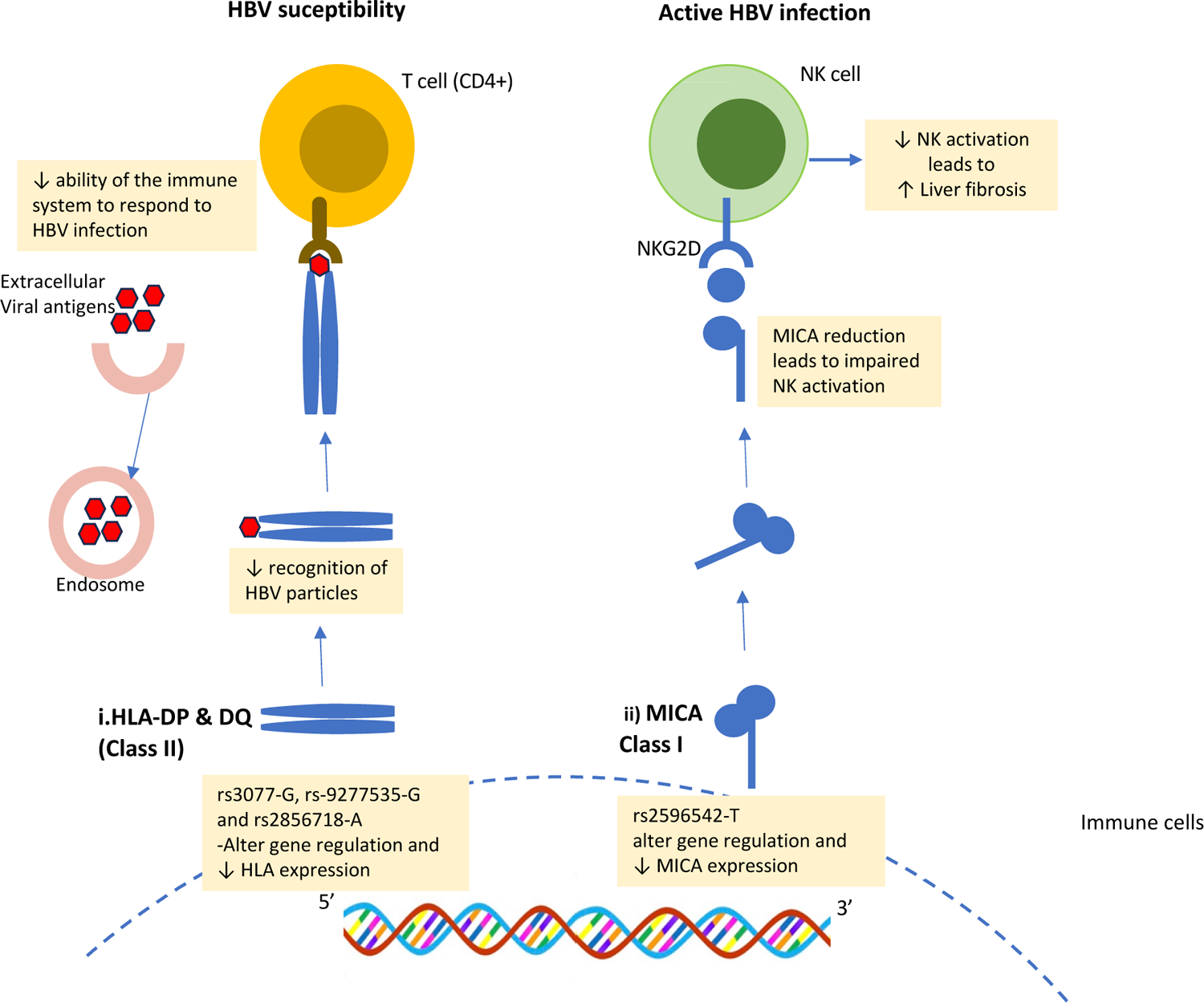
Proposed effect of genetic variations in HLA molecules on hepatitis B-associated disease. *i)* Presence of HLA-DP rs3077-G and rs9277535-G alleles and HLA-DQ rs-2856718-A allele leads to decrease expression of HLA. This results in reduce ability of HLA to recognize extracellular HBV particles and pressent to T cells. This weak response to HBV infection increases suceptibility to HBV infection. *ii)* Presence of rs2596542-T in MICA gene lead to decrease expression of MICA molecules. This results in reduce activation of NKG2D receptor found on NK cells. Since NK cells eliminate hepatic stellate cells, their impaired activation lead to increased liver fibrosis that linked to active HBV infection. HLA=Human leukocyte antigen, MICA=MHC class I Chain A, NKG2D= Natural killer group 2D, NK= Natural Killer

The same team has previously analyzed an HLA class I related SNP, rs2596542, located in the promoter region of the MHC class I chain A (MICA) gene (24). The MICA gene encodes a protein that acts as a ligand for Natural killer group 2D (NKG2D) receptors. The NKG2D is found on cytotoxic CD8, T lymphocytes, and natural killer (NK) cells, and its activation by MICA leads to the killing of infected cells and tumor cells (25). Studies have shown that MICA polymorphisms are associated with the development of Hepatitis C virus-induced HCC (26). In the Saudi cohort, a significant association of the T allele of rs2596542 was observed with the active state of HBV infection when compared with the non-active HBV group, however, there is no association of it with HCC. The SNPnexus analysis revealed that rs2596542 is located 5’ upstream in the promoter region of the MICA gene with a CADD score of 2. A study by Sharkawy et al reported that rs2596542-T allele reduces MICA gene expression in chronic hepatitis C patients and was associated with increased fibrosis stage (27). Because hepatic stellate cells play a crucial role in fibrosis progression, the low expression of MICA results in decreased NK cell activation. This reduction hampers the elimination of hepatic stellate cells by NK cells, ultimately contributing to increased fibrosis (27).

This study signifies the functional impact of rs2596542, however, this correlation has not yet been explored in HBV patients. Figure 2 illustrates the proposed effect of HLA- and MICA-SNPs on hepatitis B-associated disease.

Future studies should be designed to analyze the association of these HLA-related risk alleles with the mRNA levels of respective genes in Saudi chronic Hepatitis B patients. In addition, in vitro studies should be used to evaluate the impact of the risk alleles on immune cell function. For example, investigating the risk allele’s impact on T cells’ cytotoxic activity in response to HBV could provide valuable insights.

### Pattern recognition receptors (TLR-3, NOD2 and MD2)

Besides HLA molecules, pattern recognition receptors (PRRs) are another important part of the immune system for recognizing foreign or viral particles. Toll-like receptors (TLRs) are an example of PRRs that identify pathogens, activate intracellular antiviral pathways, and trigger the production of proinflammatory cytokines. In the context of HBV, TLR-3 recognizes viral double-stranded RNA and induces the production of interferons (IFNs) through the activation of IFN regulatory factor 3 (IRF3), which in turn inhibits HBV replication (28). In a previous study, the Al-Qahtani research group looked at nine different genetic variations of TLR3 in Saudi patients with chronic HBV infection. They found that one particular genetic variation, called rs1879026 (G/T), was significantly different between HBV carriers and uninfected controls. Additionally, the haplotype GCGA, composed of rs1879026, rs5743313, rs5743314, and rs5743315 showed a significant effect (p-value 0.034) on the susceptibility of HBV infection in persons from Saudi Arabia (29). According to SNPnexus, rs1879026 is situated in the promoter flanking region of the TLR3 gene, with a low CADD score of value 0.02, which suggests that it might not have a strong effect by itself. However, considering the result of the haplotypes and the significant role of TLR3 in immune control over HBV infection, it is still a candidate worth investigating in the future.

In a separate study, SNPs in two more PRRs were studied by the AlQahtani group, i.e., Nucleotide-binding oligomerization domain 2 (NOD2) and myeloid differentiation protein 2 (MD2) (30). NOD2 is an endosomal receptor protein expressing in macrophages, dendritic cell and heaptocytes that recognizes bacterial or viral pathogens and triggers IFN and Nuclear factor kappa B (NFkB) signaling pathways to fight infection (31). MD-2 is a co-receptor for TLR4 that contributes to pathogen recognition and production of proinflammatory cytokines (32). The investigation conducted on NOD-2 nsSNPs, rs2068844 and rs2068845, showed an association with increased risk of HBV infection or HBV susceptibility, whereas MD-2 nsSNPs, rs6472812 and rs11466004, showed an association with decreased risk of HBV infection (30). The interesting finding from SNPnexus was that both nsSNPs of NOD2 gene have deleterious effects on the structure and/or function of the protein, while SNPs of MD2 have tolerated effects on the protein (Table 1). The structural features of 1,040 amino acid long NOD2 protein, includes an N-terminal effector region critical for interaction with downstream effector proteins, a central NOD domain that governs oligomerization, and a C-terminal region responsible for ligand recognition (Figure 3a). The rs2066844-T allele is responsible for an amino acid acid change from Arginine (R) to Tryptophan (W), at position 702 in pocket 27 of the central domain, with a CADD score of 23.6. HOPE analysis showed that this substitution results in a bigger-sized amino acid residue in the resultant protein that is more hydrophobic than the wild-type residue. In addition, the charge on the mutant residue changes from positive to neutral, which could alter the folding of the protein and, in turn, affect the interaction of NOD2 with other proteins. Extensive research has shown strong association of this nsSNP with Crohn’s disease, which is a chronic inflammation of gastrointestinal tract, and the functional analysis of this SNP had shown significant reduction in NFkB signaling (33). Similarly, presence of rs2066845-C allele has been reported to reduce NOD2 ability to activate NFkB in cell culture and response to bacterial stimuli (34). In the Saudi cohort by Al-Anazi and colleagues, the homozygous CC and heterozygous GC genotypes at rs2066845 were significantly associated with HBV infection compared to the GG genotype. Additionally, the C allele significantly increased the risk of developing cirrhosis and or HCC compared to active HBV carriers (30). Supporting this, a recent study showed that hepatic NOD2 promoted HCC through proinflammatory cyokines such as TNFalpha (35). In the translated protein, the minor ‘C’ allele causes the substitution of aminoacid Glycine (Gly) to Arg at position 908. This position is within the Leucine-rich region towards the protein’s C-terminal and may have a damaging effect, reflected in a CADD score of 27.4 (Table 1). HOPE analysis predicted this substitution would result in a bigger-sized, less hydrophobic mutant residue. The wild type has no charge, but the mutation introduces a positive charge at this position, which can disturb the interaction with other molecules including ligands like viral RNA of HBV. These findings are reinforced by Mutpred2 pathogenicity score of 0.672 (from scale 0 to 1). The functional impacts include gain of helix (p value 0.02) and loss of loop (p value 0.03) in the protein by this substitution. A previous study also showed that both domains were critical for recognizing viral single-stranded RNA and activating IFN production and antiviral defense mechanisms (36). Given the functional importance of these regions, future research should prioritize biochemical assays to validate these computational predictions. Specifically, the changes in hydrophobicity and charge distribution need to be examined in vitro or in vivo to determine the involvement of mutant protein in HBV associated disease.

**Figure 3.**
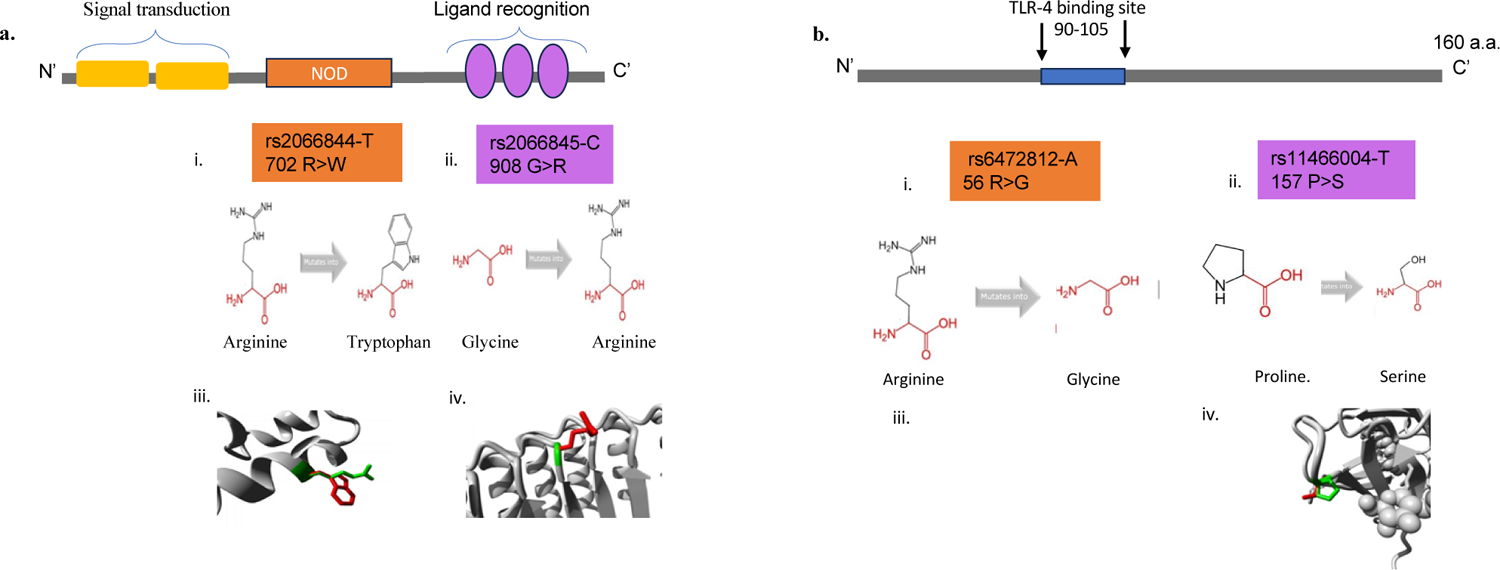
Effect of genetic variations on NOD2 and MD2 proteins structure. *a)* Schematic representation of NOD2 protein structure. i) rs-2066844-T and ii) rs-2066845-C induced amino acid substitution. Schematic structure of the original (left) and the mutant (right) amino acids. The backbone is the same for each amino acid and is colored red. The side chain unique for each amino acid is colored black. iii-iv) Closeup of the mutations by project HOPE. The protein is colored grey, the sidechain of the wild-type residue is presented as green, and the mutant residue is shown in red. Schematic representation of MD2 protein structure. i) rs6472812-A and ii) rs-11466004 induced amino acid substitution. Schematic structure of the original (left) and the mutant (right) amino acids.

For MD2, also known as Lymphocyte antigen 96 (LY96), the risk or minor alleles i.e. rs6472812-A and rs11466004-T, have decreased frequencies in HBV patients than in controls (A= 21.2% versus 38.9% and T= 23.5% versus 41.8%). The authors conclude that these alleles may reduce the risk of HBV infection and could play a protective role against the development of HBV infection (30). The authors also reported these SNPs to be in the intronic region, however, per SNPnexus, SNP rs6472812 and rs11466004 are located in the 2nd and 5th exonic (coding) region of the MD2 gene. The possible explanation for this discrepancy could be the presence of multiple transcript variants encoding different isoforms of MD-2 that might be missing the exon with specific SNPs. MD2 is a 160 a.a. long protein that binds to the extracellular domain of TLR-4 (37). The Hope analysis was unsuccessful for this mutation after several attempts, hence, we used ProtVar (38) for additional annotations. According to ProtVar rs6472812-A leads to substituting Arg to Gly at position 56 near N-terminal (Figure 3b), which is more likely to be benign, with a CADD score of 11.2. Additionally, this position is predicted to interact with IRF-3 protein with a confidence score of 0.369 that is considered as high model confidence. A previous crystallography study by Chen et al has shown that this mutation site is near cysteine 51 which is important for the structural stability of the protein (37). For rs1146004, the presence of the T allele led to substituting a.a. Proline (Pro) to Serine (Ser) at position 157, resulting in a protein that is also benign or tolerated. The predicted changes in protein by HOPE include a smaller residue with less hydrophobicity that could impact its interaction with other molecules.

The backbone is the same for each amino acid and is colored red. The side chain unique for each amino acid is colored black. iii-iv) Closeup of the mutations by project HOPE. The protein is colored grey, the sidechain of the wild-type residue is presented as green, and the mutant residue is shown in red.

Literature search provide evidence that TLR4 regulates several major inflammatory signaling pathways including transcription factors, NFkB and IRF3 (28, 39). Since MD2 is a co-receptor for TLR4, its favorable SNPs against hepatitis B could enhance the TLR-4 mediated recognition of viral particles, which could lead to increase production of cytokines that recruit immune cells to the site of infection and promote clearance of infected cells. This hypothesis can be investigated in vitro or in vivo by examining the immune cell recruitment and HBV clearance. Wang et al reported that activation of the TLR4 signaling pathway induces increased production of neutrophils and promotes the immune response and clearance of HBV (40). Furthermore, investigating the association of the risk allele with HBV treatment outcome will also give insights into the favorable impact of these SNPs. Figure 4 illustrates the proposed effect of a) NOD2 and b) MD2 SNPs on HBV suceptibility and clearance.

**Figure 4.**
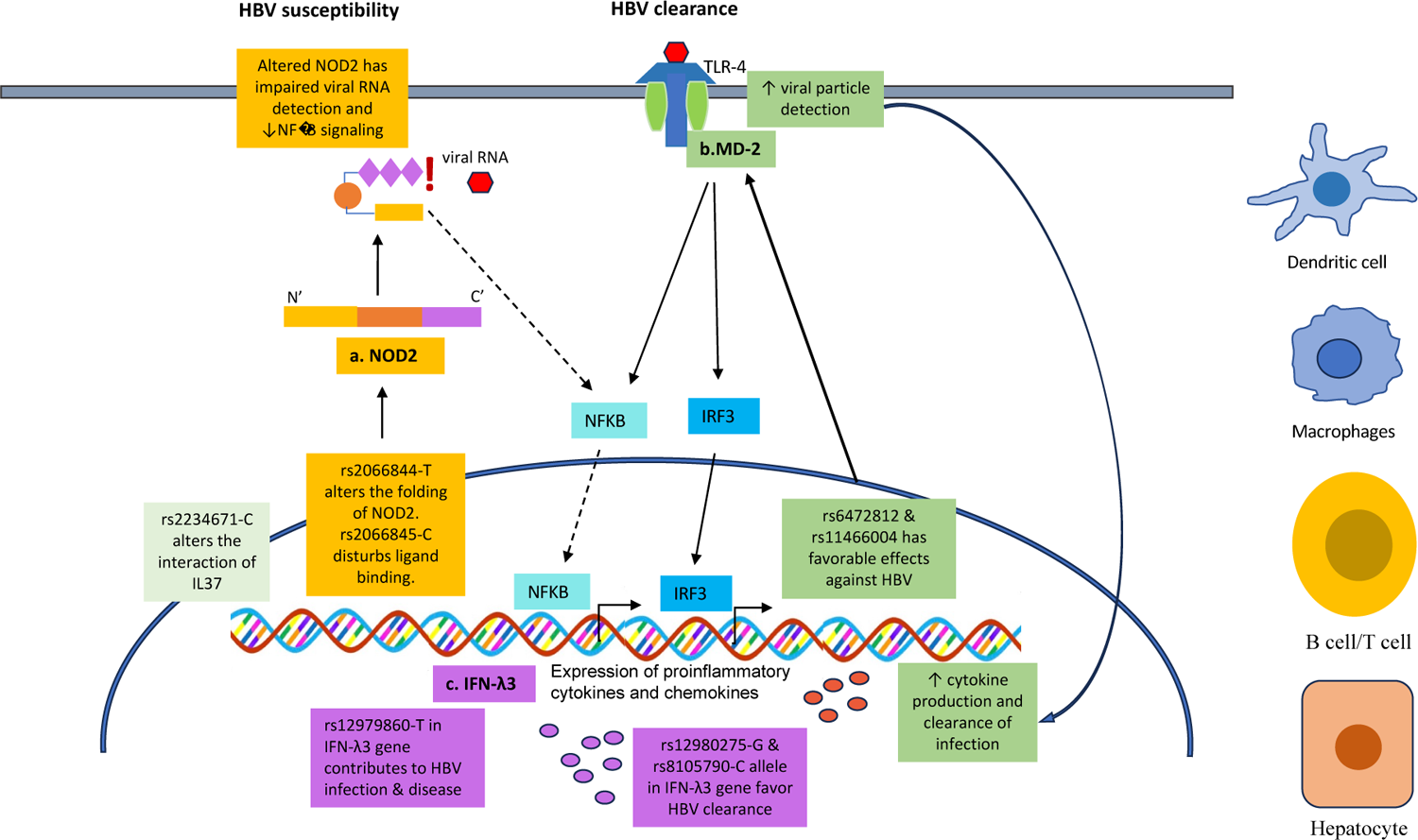
Proposed effect of genetic variations in NOD2, MD2, IL28 and IL37 on hepatitis B-associated disease. *a)* Presence of rs2068844-T and rs2066845 alleles in NOD-2 gene modulate the folding and ligand binding ability of the protein. This results in reduce ability to recognize viral RNA and decrease NFKB siganling. This weak response to HBV infection increases suceptibility to HBV infection. *b)* MD2 is a co-receptor for TLR-4. The rs6472812-A and rs11466004-T alleles in MD2 gene lead to enhanced activation of transcriotion factors such as NFKB and IRF3.This results in increased production of cytokines and clearance of HBV infection. *c)* IL28B or IFN-λ3 NOD2=Nucleotide binding oligomerization domain 2, MD2= myeloid differentiation protein 2, NFκB = Nuclear factor Kappa B, IRF = Interferon regulatory factor 3, TLR-4 = Toll like receptor.

### Chemokine receptor (CXCR1)

The chemokine (C-X-C motif) receptor 1 (CXCR1) is a G-protein-coupled receptor, expressed predominantly on neutrophils and binds to interleukin-8 (IL-8), which is a pro-inflammatory chemokine. IL-8/CXCR1 binding activates and migrates neutrophils, T cells, and basophils to the site of inflammation, and hence plays a key role in inflammatory disorders including hepatic inflammatory response (41). A study in the Saudi population by Almajhdi et al found that SNP rs2234671 in the coding region of the CXCR1 gene is significantly associated with the active state of chronic HBV infection when compared with the inactive group (42). Nevertheless, no correlation was found between CXCR1 risk allele and HBV-mediated cirrhosis and HCC. The interesting finding was that in the Saudi population, the C allele was the minor allele while in the other populations’ data G allele is reported to be the minor allele and its presence leads to substituting the amino acid Ser to Threonine (Thr) at position 276 in the protein. This situation will be the opposite in the HBV-associated Saudi population, where the presence of rs2234671-C will lead to substituting Thr with Ser. The structural analysis by HOPE showed that Ser is a smaller residue than Thr, and substitution resulted in a tolerated effect with a score of 2.9 which indictes that while the substitution may influence protein interactions, it is unlikely to significantly contribute to pathogenic conditions. A study by Xu and colleagues showed low expression of CXCR1 on neutrophils from chronic hepatitis B patients compared to healthy controls (43). However, this was not checked in the Saudi cohort, hence, the effect of this SNP on the corresponding protein is not known. Infact up till now there is limited data on CXCR1 role in HBV infection. Further exploring the expression of CXCR1 in neutrophils from patients with risk alleles and correlate it with the clinical parameters of HBV disease severity might yield valuable insights.

### Cytokines

In addition to examining SNPs in HLA and PRR molecules, the Al-Qahtani research group investigated SNPs in a few specific cytokines namely Interleukin-28B and Interleukin-37 among the Saudi population (44, 45). Cytokines are small signaling proteins released by immune cells in response to infections. They play a crucial role in monitoring HBV infection and targeting different steps of its life cycle (46).

#### Interleukin 28B (IL28B) or Interferon lambda 3 (IFN-λ3)

Interleukin-28B (IL28B) gene encodes for a cytokine interferon lambda 3 (IFN-λ3), which has antiviral and immunomodulatory properties. Genetic variations in IL28B have been widely researched in viral hepatitis, especially hepatitis C, because of its role in predicting treatment outcomes and guiding therapeutic decisions (47). In the case of HBV, its role is less clear but research showed that certain IL-28B polymorphisms may affect spontaneous clearance of the virus and influence the response to Pegylated IFN-alpha therapy (48). The Al-Qahtani group studied five IL28B SNPs in HBV-infected patients with the clearance group as control. Their results revealed that rs12980275 and rs8105790 were significantly associated with HBV clearance. The frequency of their risk alleles, i.e., rs12980275-G and rs8105790-C, were more prevalent in individuals who cleared HBV compared to chronic HBV carriers, which the authors concluded to be protective against HBV infection (45). However, they did not assess the impact of these SNPs on treatment response in HBV-infected patients. According to SNPnexus, these SNPs are located 3’ downstream of the IFN-λ3 gene with very low CADD scores, and their predicted function is related to Histones (Table 2). The low CADD score support the idea that these SNPs donot contribute to disease development rather they may facilitate viral clearance by enhancing IFN-λ3 function and strengthening the host’s antiviral response. Another significnat finding from the study of Al-Qahtani’s group was the association of rs12979860-T allele with susceptibility to HBV as well as active infection, including cirrhosis and HCC (Table 2). SNPNexus revealed that rs12979860 is located upstream of the IL28B promoter region at a CpG (cytosine followed by Guanosine) site, which can regulate gene expression via methylation (49). A study involving Saudi patients with HCV found that presence of the rs12979860-CC genotype was linked to better treatment response while the presence of the TT genotype was associated with poor treatment outcomes (50), however, no data related to Hepatitits B patients is available. Another research showed that rs12979860-T reduces the expression of IFN-λ3 in the liver and peripheral blood mononuclear cells of hepatitis C patients (51). These findings suggest that the presence of the T allele may lower IFN-λ3 expression, contributing to chronic HBV infection and active disease as depicted in figure 4c. As no research has examined these connections in the Saudi population, additional studies are required to explore how these identified SNPs affect treatment outcomes with IFN in HBV patients. Multicenter investigations are crucial for validating and expanding these results for clinical use.

#### Interleukin 37 (IL37)

Interleukin 37 (IL37) exhibits broad anti-inflammatory and immunomodulatory effects, functioning in both intracellular and extracellular forms. In response to inflammatory stimuli, the intracellular IL37 suppresses gene expression of proinflammatory cytokines by translocating into the nucleus, whereas the extracellular form binds to IL18 receptor α (IL18Rα) and inhibits proinflammatory pathways such as NFκB and MAPK (52). Although the role of IL37 in HBV pathogenesis is not fully understood, considerable evidence suggests that in response to liver injury, IL37 can downregulate several proinflammatory cytokines while upregulating antiinflammatory factors, thus potentially influencing HBV infection (53). A total of ten SNPs were tested in the cohort of the Saudi population, out of which three SNPs, rs2723175, rs2708973 and rs28947200, were found associated with a high risk of HBV infection (44). SNPnexus showed that rs2708973 has a CADD score of 4.56, and its predicted function is categorized as an enhancer, signifying its important role in the regulation of IL37 gene. A previous GWAS in HIV-infected patients has shown the association of this SNP with retroviral therapy (54), however, in the context of HBV, there is a lack of data regarding its association with HBV treatment outcomes and warrants further investigation. The only non-synonymous SNP in their study, rs28947200, showed T allele association with HBV persistence when compared to the clearance group. Further computational analysis revealed that the presence of minor T allele results in a substitution of Arginine (R) to Tryptophan (W) at position 152 in the 218 amino acid long IL37 protein. This amino acid change introduces a larger, more hydrophobic residue compared to the wild type and with a neutral charge. According to HUPO report the mutated residue is located in a domain that is important for binding to other molecules and alter protein function. In addition, wildtype redisue at this position form a salt bridge with glutamic acid at position 148 but the difference in charge will disturb the ionic interaction made by the original wild-type residue. The CADD score for this substitution is 19.1, suggesting a possibly damaging effect. This may affect the protein’s ability to modulate the immune response effectively and contribute to HBV persistence.

Conversely, four SNPs—rs2723176, rs2723186-AG, rs4364030-G, and rs4392270-A—were significantly linked to the clearance of HBV infection. This suggests that their presence either upregulates IL37 expression or enhances its effectiveness in clearing HBV infection. A recent study has suggested that IL37 offers protection against hepatitis and liver damage by suppressing the cytotoxic effects of CD8+ T cells activated by HBV peptides (55). Overall, these findings suggest that specific genetic variants in IL37 gene can influence both the risk of HBV infection and the subsequent immune response, leading to varied outcomes in disease persistence or clearance. To further elucidate these relationships, the intrahepatic expression of IL37 should be evaluated in different stages of hepatitis B as well as the cytokine profile affected by IL37.

### Conclusion and Directions for Future Research

Extensive research has consistently demonstrated that SNPs within host genes have the potential to influence protein expression, ultimately playing a role in determining an individual’s vulnerability to diseases. By investigating SNPs, researchers can identify individuals predisposed to certain diseases, leading to improved screening, diagnosis, and treatment decisions (56). One powerful illustration of the significance of SNP analysis lies in the use of BRCA gene mutations in the early diagnosis and treatment of breast cancer (57). Hepatitis B virus infection and its mediated liver complications remain a long-standing public health challenge worldwide. Over the past decade, genetic studies on HBV-infected individuals in the Saudi population have identified a series of SNPs significantly associated with either protection against HBV, susceptibility to HBV or increased severity of HBV-mediated disease. Notably, these SNPs have not been thoroughly explored or incorporated into follow-up research. This review used computational prediction tools to assess the functional roles of these SNPs and their impact on host defense and HBV susceptibility, that could in turn help to prioritize them for further analysis.

The results identified rs3077 and rs-2856718 in HLA-DP and DQ genes to have the highest CADD score among the non-coding variants that were related to HBV susceptibility. Their predicted function as promoter or histone-related regulatory elements influence HLA expression in the antigen presenting cells and thereby affects immune recognition and response to HBV infection. Among the coding SNPs, the NOD2 gene mutations, rs2068844 and rs2066845 were predicted as deleterious ones with possible structural changes in protein structure. Presence of risk alleles is predicted to disrupt ligand binding and protein folding that could affect the activation of transcription factors such as NFKB and IRF, responsible for the production of proinflammatory cytokines, and thus facilitate HBV infection. Notably, rs 2066845 turned out to have the highest pathogenecity score of all the nsSNPs. NOD2 has gained significant attention in recent years due to its role in inflammation and hepatitis, which subsequently contribute to the development of HCC and highlight its potential as a therapeutic target (58). This emphasizes the importance of further analysis of this SNP in HBV infection and clinical outcomes. The SNPs data from the cytokines IL28B and IL37 also suggest a complex relationship between these gene and HBV infection dynamics within the Saudi population. In addition, the coding SNPs in MD2 gene were associated with the clearance of HBV infection and need further investigation to explore the potential as biomarkers.

Despite these significant findings, there were some critical limitations in these studies, such as none of the studies have addressed the relationship between SNPs and HBV treatment outcomes, which restricts the clinical implication of these genetic variations. To address this limitation, the SNPs found associated with HBV clearance (Table 2) should be further analyzed to investigate their potential to predict treatment response in HBV patients. Additionally, the SNPs associated with active HBV status (i.e. FDX1, MICA, NOD2, CXCR1, IL28B and IL37) should be validated on large-scale to identify their potential as biomarkers to predict HBV progression. Developing these SNPs as biomarkers for HBV susceptibility, progression, and treatment response will also require multicentre validation studies. Furthermore, integrating all related SNPs in a multiomics approach, combining genomic, transcriptomic, and proteomic data, will enhance our understanding of the complex interactions involved in the immune response to HBV at various stages of infection.

By integrating these research approaches, the findings on HBV-associated SNPs can progress from genetic associations to practical clinical applications.This will not only enhance precision medicine strategies for HBV management but also contribute to reducing disease prevalence and improving patient outcomes in Saudi Arabia and globally.

## Data Availability

All data produced in the present work are contained in the manuscript

---

**SuppTable 1.**
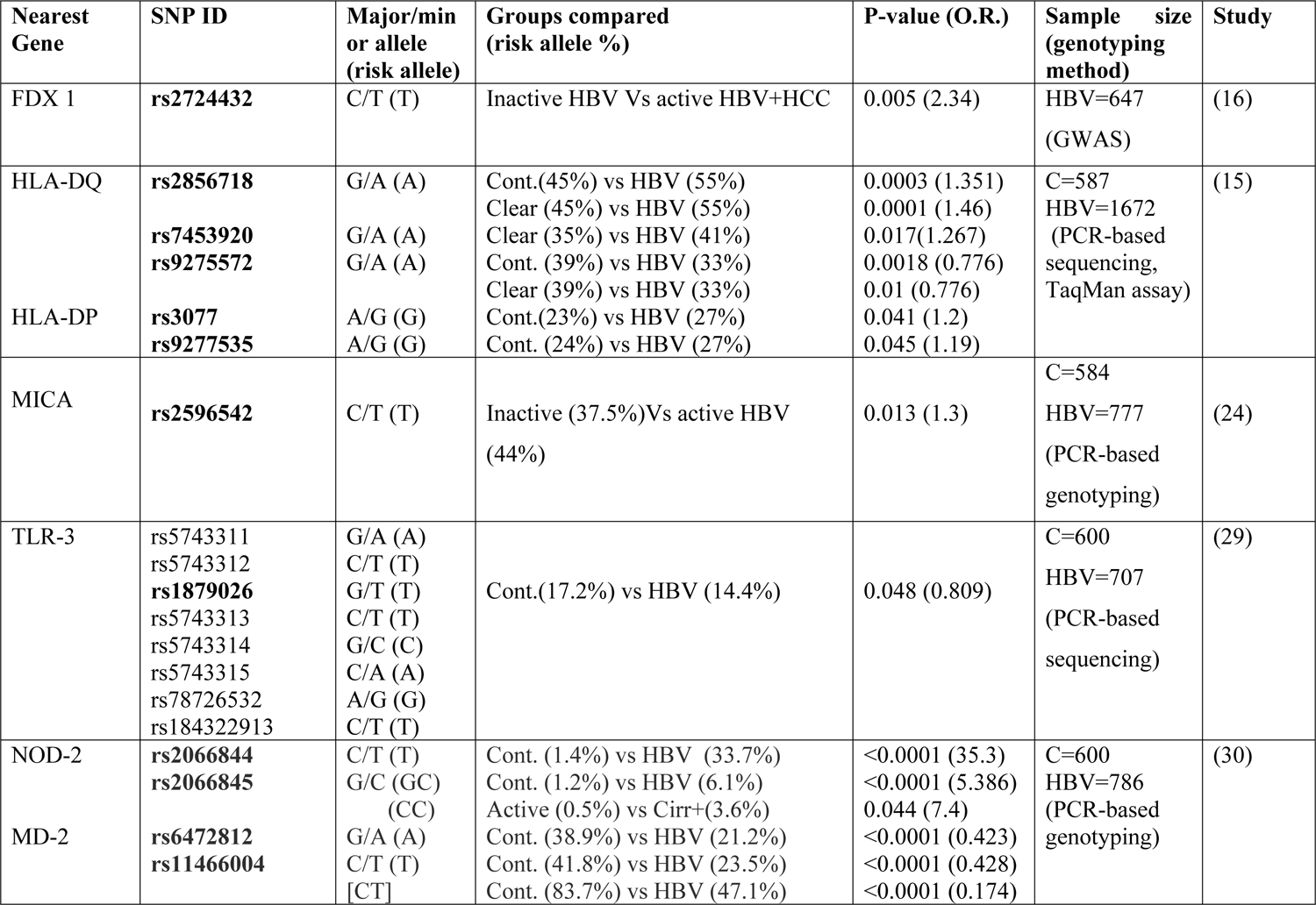
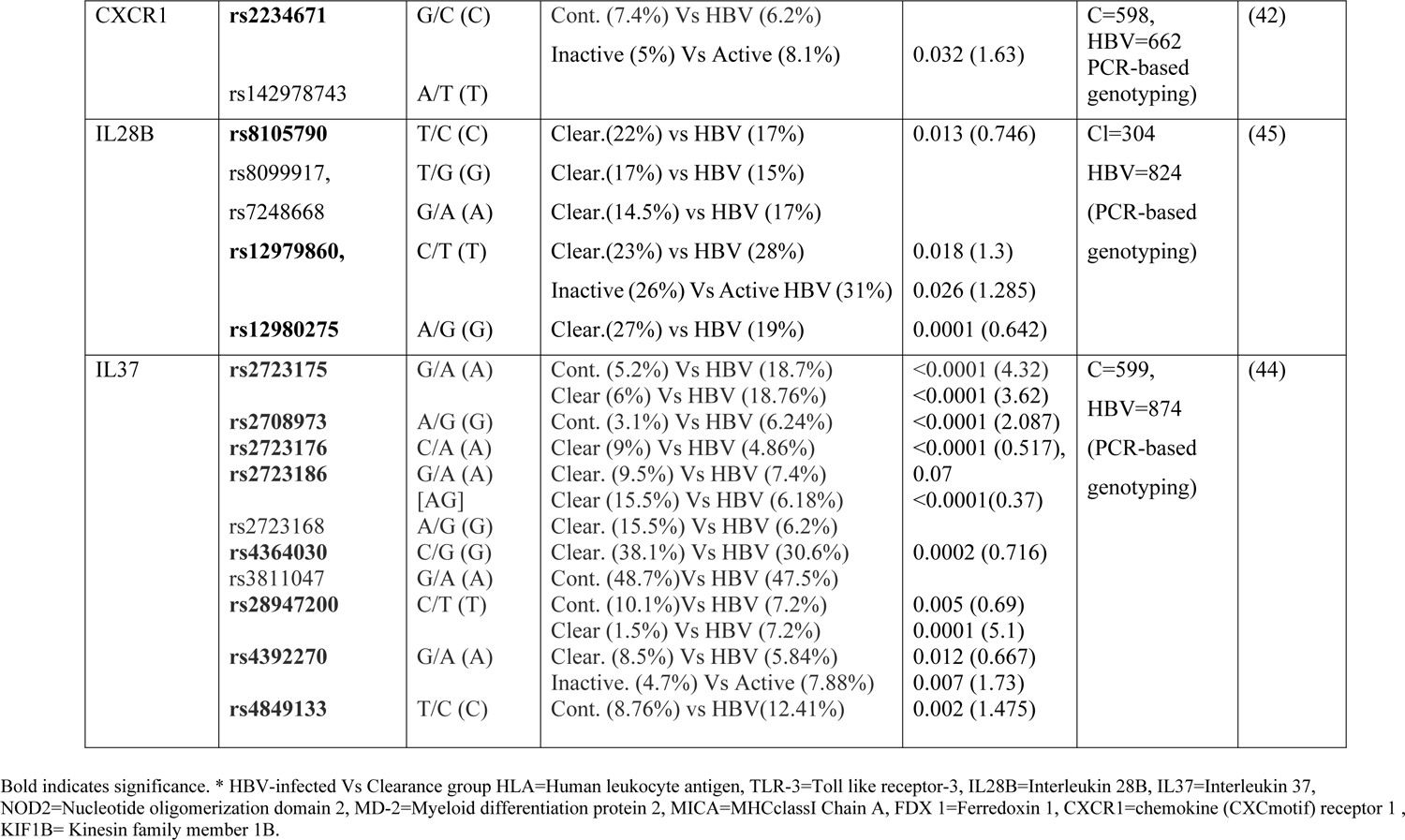
Gene polymorphisms associated with hepatitis B virus infection/clearance in Saudi population.

## Notes

### Competing Interest Statement

The authors have declared no competing interest.

### Funding Statement

This study did not receive any funding

### Author Declarations

The study used ONLY openly available human data that were originally located at PUBMED or NCBI websites.

## References

1. World Health Organization W. Hepatitis C fact sheet: https://www.who.int/news-room/fact-sheets/detail/hepatitis-c; 2021 [Available from:.

2. Fattovich G, Bortolotti F, Donato F. Natural history of chronic hepatitis B: special emphasis on disease progression and prognostic factors. J Hepatol. 2008;48(2):335–52.

3. Alghamdi M, Alghamdi AS, Aljedai A, Khathlan AA, Masri NA, Qutub A, et al. Revealing Hepatitis B Virus as a Silent Killer: A Call-to-Action for Saudi Arabia. Cureus. 2021;13(5):e14811.

4. Sanai FM, Alghamdi H, Alswat KA, Babatin MA, Ismail MH, Alhamoudi WK, et al. Greater prevalence of comorbidities with increasing age: Cross-sectional analysis of chronic hepatitis B patients in Saudi Arabia. Saudi J Gastroenterol. 2019;25(3):194–200.

5. Sanai F, Alkhatry M, Alzanbagi A, Kumar S. Hepatitis B virus infection in Saudi Arabia and the UAE: Public health challenges and their remedial measures. J Infect Public Health. 2023;16(9):1410–7.

6. Rybicka M, Woziwodzka A, Romanowski T, Sznarkowska A, Stalke P, Dreczewski M, et al. Host genetic background affects the course of infection and treatment response in patients with chronic hepatitis B. J Clin Virol. 2019;120:1–5.

7. Yang F, Wu L, Xu W, Liu Y, Zhen L, Ning G, et al. Diverse Effects of the NTCP p.Ser267Phe Variant on Disease Progression During Chronic HBV Infection and on HBV preS1 Variability. Front Cell Infect Microbiol. 2019;9:18.

8. Shastry BS. SNPs: impact on gene function and phenotype. Methods Mol Biol. 2009;578:3–22.

9. Cargill M, Altshuler D, Ireland J, Sklar P, Ardlie K, Patil N, et al. Characterization of single-nucleotide polymorphisms in coding regions of human genes. Nat Genet. 1999;22(3):231–8.

10. Manolio TA. Genomewide association studies and assessment of the risk of disease. N Engl J Med. 2010;363(2):166–76.

11. Zhang Z, Wang C, Liu Z, Zou G, Li J, Lu M. Host Genetic Determinants of Hepatitis B Virus Infection. Front Genet. 2019;10:696.

12. Xu J, Zhan Q, Fan Y, Yu Y, Zeng Z. Human genetic susceptibility to hepatitis B virus infection. Infect Genet Evol. 2021;87:104663.

13. Oscanoa J, Sivapalan L, Gadaleta E, Dayem Ullah AZ, Lemoine NR, Chelala C. SNPnexus: a web server for functional annotation of human genome sequence variation (2020 update). Nucleic Acids Res. 2020;48(W1):W185–W92.

14. Rentzsch P, Witten D, Cooper GM, Shendure J, Kircher M. CADD: predicting the deleteriousness of variants throughout the human genome. Nucleic Acids Res. 2019;47(D1):D886–D94.

15. Al-Qahtani AA, Al-Anazi MR, Abdo AA, Sanai FM, Al-Hamoudi W, Alswat KA, et al. Association between HLA variations and chronic hepatitis B virus infection in Saudi Arabian patients. PLoS One. 2014;9(1):e80445.

16. Al-Qahtani A, Khalak HG, Alkuraya FS, Al-hamoudi W, Alswat K, Al Balwi MA, et al. Genome-wide association study of chronic hepatitis B virus infection reveals a novel candidate risk allele on 11q22.3. J Med Genet. 2013;50(11):725–32.

17. Sheftel AD, Stehling O, Pierik AJ, Elsässer HP, Mühlenhoff U, Webert H, et al. Humans possess two mitochondrial ferredoxins, Fdx1 and Fdx2, with distinct roles in steroidogenesis, heme, and Fe/S cluster biosynthesis. Proc Natl Acad Sci U S A. 2010;107(26):11775–80.

18. Sun B, Ding P, Song Y, Zhou J, Chen X, Peng C, et al. FDX1 downregulation activates mitophagy and the PI3K/AKT signaling pathway to promote hepatocellular carcinoma progression by inducing ROS production. Redox Biol. 2024;75:103302.

19. Biterge B, Schneider R. Histone variants: key players of chromatin. Cell Tissue Res. 2014;356(3):457–66.

20. Traherne JA. Human MHC architecture and evolution: implications for disease association studies. Int J Immunogenet. 2008;35(3):179–92.

21. Deng N, Zhou H, Fan H, Yuan Y. Single nucleotide polymorphisms and cancer susceptibility. Oncotarget. 2017;8(66):110635–49.

22. Hong D, Jeong S. 3’UTR Diversity: Expanding Repertoire of RNA Alterations in Human mRNAs. Mol Cells. 2023;46(1):48–56.

23. O’Brien TR, Kohaar I, Pfeiffer RM, Maeder D, Yeager M, Schadt EE, et al. Risk alleles for chronic hepatitis B are associated with decreased mRNA expression of HLA-DPA1 and HLA-DPB1 in normal human liver. Genes Immun. 2011;12(6):428–33.

24. Al-Qahtani AA, Al-Anazi M, Abdo AA, Sanai FM, Al-Hamoudi W, Alswat KA, et al. Genetic variation at −1878 (rs2596542) in MICA gene region is associated with chronic hepatitis B virus infection in Saudi Arabian patients. Exp Mol Pathol. 2013;95(3):255–8.

25. Carapito R, Bahram S. Genetics, genomics, and evolutionary biology of NKG2D ligands. Immunol Rev. 2015;267(1):88–116.

26. Luo X, Wang Y, Shen A, Deng H, Ye M. Relationship between the rs2596542 polymorphism in the MICA gene promoter and HBV/HCV infection-induced hepatocellular carcinoma: a meta-analysis. BMC Med Genet. 2019;20(1):142.

27. Sharkawy RE, Bayoumi A, Metwally M, Mangia A, Berg T, Romero-Gomez M, et al. A variant in the MICA gene is associated with liver fibrosis progression in chronic hepatitis C through TGF-β1 dependent mechanisms. Sci Rep. 2019;9(1):1439.

28. Zhang E, Lu M. Toll-like receptor (TLR)-mediated innate immune responses in the control of hepatitis B virus (HBV) infection. Med Microbiol Immunol. 2015;204(1):11–20.

29. Al-Qahtani A, Al-Ahdal M, Abdo A, Sanai F, Al-Anazi M, Khalaf N, et al. Toll-like receptor 3 polymorphism and its association with hepatitis B virus infection in Saudi Arabian patients. J Med Virol. 2012;84(9):1353–9.

30. Al-Anazi MR, Nazir N, Abdo AA, Sanai FM, Alkahtani S, Alarifi S, et al. Genetic variations of NOD2 and MD2 genes in hepatitis B virus infection. Saudi J Biol Sci. 2019;26(2):270–80.

31. Moreira LO, Zamboni DS. NOD1 and NOD2 Signaling in Infection and Inflammation. Front Immunol. 2012;3:328.

32. Shimazu R, Akashi S, Ogata H, Nagai Y, Fukudome K, Miyake K, et al. MD-2, a molecule that confers lipopolysaccharide responsiveness on Toll-like receptor 4. J Exp Med. 1999;189(11):1777–82.

33. Parkhouse R, Monie TP. Dysfunctional Crohn’s Disease-Associated NOD2 Polymorphisms Cannot be Reliably Predicted on the Basis of RIPK2 Binding or Membrane Association. Front Immunol. 2015;6:521.

34. Bonen DK, Ogura Y, Nicolae DL, Inohara N, Saab L, Tanabe T, et al. Crohn’s disease-associated NOD2 variants share a signaling defect in response to lipopolysaccharide and peptidoglycan. Gastroenterology. 2003;124(1):140–6.

35. Zhou Y, Hu L, Tang W, Li D, Ma L, Liu H, et al. Hepatic NOD2 promotes hepatocarcinogenesis via a RIP2-mediated proinflammatory response and a novel nuclear autophagy-mediated DNA damage mechanism. J Hematol Oncol. 2021;14(1):9.

36. Sabbah A, Chang TH, Harnack R, Frohlich V, Tominaga K, Dube PH, et al. Activation of innate immune antiviral responses by Nod2. Nat Immunol. 2009;10(10):1073–80.

37. Chen L, Fu W, Zheng L, Wang Y, Liang G. Recent progress in the discovery of myeloid differentiation 2 (MD2) modulators for inflammatory diseases. Drug Discov Today. 2018;23(6):1187–202.

38. Stephenson JD, Totoo P, Burke DF, Jänes J, Beltrao P, Martin MJ. ProtVar: mapping and contextualizing human missense variation. Nucleic Acids Res. 2024;52(W1):W140–W7.

39. Akira S, Takeda K. Toll-like receptor signalling. Nat Rev Immunol. 2004;4(7):499–511.

40. Wang Q, Zhang H, Chen Z, Chen L, Pan F, Zhou Q. Proliferation of CD11b+ myeloid cells induced by TLR4 signaling promotes hepatitis B virus clearance. Cytokine. 2022;153:155867.

41. Holmes WE, Lee J, Kuang WJ, Rice GC, Wood WI. Structure and functional expression of a human interleukin-8 receptor. Science. 1991;253(5025):1278–80.

42. Almajhdi FN, Al-Ahdal M, Abdo AA, Sanai FM, Al-Anazi M, Khalaf N, et al. Single nucleotide polymorphisms in CXCR1 gene and its association with hepatitis B infected patients in Saudi Arabia. Ann Hepatol. 2013;12(2):220–7.

43. Xu R, Bao C, Huang H, Lin F, Yuan Y, Wang S, et al. Low expression of CXCR1/2 on neutrophils predicts poor survival in patients with hepatitis B virus-related acute-on-chronic liver failure. Sci Rep. 2016;6:38714.

44. Al-Anazi MR, Matou-Nasri S, Al-Qahtani AA, Alghamdi J, Abdo AA, Sanai FM, et al. Association between IL-37 gene polymorphisms and risk of HBV-related liver disease in a Saudi Arabian population. Sci Rep. 2019;9(1):7123.

45. Al-Qahtani AA, Al-Anazi MR, Abdo AA, Sanai FM, Al-Hamoudi WK, Alswat KA, et al. Genetic variation in interleukin 28B and correlation with chronic hepatitis B virus infection in Saudi Arabian patients. Liver Int. 2014;34(7):e208–16.

46. Xia Y, Protzer U. Control of Hepatitis B Virus by Cytokines. Viruses. 2017;9(1).

47. Ge D, Fellay J, Thompson AJ, Simon JS, Shianna KV, Urban TJ, et al. Genetic variation in IL28B predicts hepatitis C treatment-induced viral clearance. Nature. 2009;461(7262):399–401.

48. Ying SY, Hu YR, Gao GS, Lou KH, Huang Z. Polymorphisms Predict the Efficacy of Peginterferon Alpha in Patients With Chronic Hepatitis B: A Meta-Analysis. Front Med (Lausanne). 2021;8:691365.

49. Robertson KD. DNA methylation and human disease. Nat Rev Genet. 2005;6(8):597–610.

50. Abdo AA, Al-Ahdal MN, Khalid SS, Helmy A, Sanai FM, Alswat K, et al. IL28B polymorphisms predict the virological response to standard therapy in patients with chronic hepatitis C virus genotype 4 infection. Hepatol Int. 2013;7(2):533–8.

51. Langhans B, Kupfer B, Braunschweiger I, Arndt S, Schulte W, Nischalke HD, et al. Interferon-lambda serum levels in hepatitis C. J Hepatol. 2011;54(5):859–65.

52. Su Z, Tao X. Current Understanding of IL-37 in Human Health and Disease. Front Immunol. 2021;12:696605.

53. Jiang YG, Wang YM, Liu TH, Liu J. Association between HLA class II gene and susceptibility or resistance to chronic hepatitis B. World J Gastroenterol. 2003;9(10):2221–5.

54. Leger PD, Johnson DH, Robbins GK, Shafer RW, Clifford DB, Li J, et al. Genome-wide association study of peripheral neuropathy with D-drug-containing regimens in AIDS Clinical Trials Group protocol 384. J Neurovirol. 2014;20(3):304–8.

55. Liu Q, Zhou Q, Wang M, Pang B. Interleukin-37 suppresses the cytotoxicity of hepatitis B virus peptides-induced CD8+ T cells in patients with acute hepatitis B. Biomol Biomed. 2023;23(3):527–34.

56. Meyerson M. Human genetic variation and disease. Lancet. 2003;362(9380):259–60.

57. van Veen EM, Brentnall AR, Byers H, Harkness EF, Astley SM, Sampson S, et al. Use of Single-Nucleotide Polymorphisms and Mammographic Density Plus Classic Risk Factors for Breast Cancer Risk Prediction. JAMA Oncol. 2018;4(4):476–82.

58. Omaru N, Watanabe T, Kamata K, Minaga K, Kudo M. Activation of NOD1 and NOD2 in the development of liver injury and cancer. Front Immunol. 2022;13:1004439.

